# Trends in age, sex and racial differences in the incidence of infective endocarditis in Florida and New York

**DOI:** 10.1101/2023.05.17.23290144

**Authors:** Anderson Anuforo, Ehimen Aneni, Emmanuel Akintoye, Nnabuchi Anikpezie, Smit D. Patel, Ayorinde Soipe, Eloho Olojakpoke, Devin Burke, Julius Gene Latorre, Priyank Khandelwal, Seemant Chaturvedi, Bruce Ovbiagele, Fadar Oliver Otite

**Author notes:** Correspondence: Fadar Oliver Otite; Cerebrovascular and Neurocritical Care Division, Upstate Neurological Institute, Syracuse, New York 750 East Adams St, Syracuse, NY 13210.

## Abstract

**Background:** How the incidence of infective endocarditis (IE) changed in various age, sex and racial/ethnic subgroups of the United States along with the worsening opioid epidemic over the last decade is unknown.

**Methods:** We utilized data from the 2007-2018 State Inpatient Databases (SID)) of two large demographically diverse states (Florida and New York) to conduct a retrospective cohort study. Cases of incident IE identified using validated International Classification of Diseases codes were combined with census data to compute age, sex- and race-specific incidence. Joinpoint regression was used to quantify the annualized percentage change (APC) in incidence over time.

**Results:** Of 98,221 incident IE admissions, 70.0% were Non-Hispanic White (NHW) and 60.6% were ≥ 65 years old (yo). The average annual age and sex-standardized incidence of IE in cases/100,000 population was 19.2 (95%CI 18.7-19.6) but this varied by age, sex and race. Incidence was ≈20% higher in men (20.9 [95%CI 20.2-21.7]) compared to women (17.5 [95%CI 16.8-18.2]) and increased with age in both sexes. Incidence was higher in Non-Hispanic Blacks (NHB); 22.5(95%CI 21.0-23.9) compared to NHW: 20.0(95%CI 19.3 to 20.6), Hispanic: 13.2(95%CI 12.3-14.1) and Asian/Pacific Islander patients: 5.9(95%CI 4.7 to 7.1). The age and sex-standardized incidence did not change over time (APC 0.5%, *p=*0.646). However, incidence increased in women 18-44 (APC 11.0%, *p<*0.001), men 18-44 (APC 7.3, *p<*0.001) and 45-64 yo (APC 1.5%, *p=*0.002) but declined in women ≥ 65 yo (APC −2.8, *p=*0.049). Most of this increased incidence occurred in NHW women 18-44 (APC 16.6%, *p<*0.001), NHW men 18-44 (APC 10.6%, *p<*0.001), NHW men 45-64 (APC 2.8%, *p<*0.001) and Hispanic Men 18-44 yo (APC 5.6%, *p<*0.001). Incidence did not change over time in these age/sex groups of NHB and in other age/sex groups. Prevalence of opioid use disorder increased by >2-fold in all age/sex groups over time but the pace of increase was faster in NHW women and men 18-44 yo compared to other races of similar demography (p-for-time interaction <0.001).

**Conclusion:** Over the last two decades, among residents of Florida and New York, incidence of IE increased in several demographic groups, but the most prominent rise was among young NHWs, particularly young NHW women.

## Introduction

Infective endocarditis (IE) is a rare but severe infection of the cardiovascular system that is associated with significant mortality.^1^ IE risk increases with age^2^ and disproportionately affects men compared to women^3^, but the epidemiological characteristics of IE in the United States (US) may be changing. Over the last two decades, the opioid epidemic has exponentially worsened the IE burden amongst the predominantly young persons who inject drugs (PWID)^4^. Life expectancy of patients with congenital heart diseases who carry an exaggerated risk of IE is improving^5^ and more patients with multiple comorbidities including those with end-stage renal disease on hemodialysis now live longer^6,7^. Moreover, the US population is aging^8^ and the utilization of implantable cardiac prosthetics and devices, mainly in older adults with multiple comorbidities, has also increased^9^. Consequent to their varying prevalence in various age, sex and racial/ethnic subgroups of the population, all these aforementioned factors may have led to divergent shifts in the demographic characteristics of the population at highest risk for IE. However, data on IE incidence trends in various demographic subgroups of the US are lacking.

The primary aims of this study are to (1) quantify race-, age-, and sex-specific incidence of IE in the US and (2) to investigate trends in the incidence of IE in various racial subgroups of men and women of the US over the last decade.

## Methods

We used the 2007-2018 State Inpatient Databases of Florida and New York to conduct a retrospective cohort study. The SIDs are a family of databases by the Health Care Utilization Project (HCUP) of the Agency for Healthcare Research and Quality that contain all acute inpatient discharges in participating states. The two selected states are large demographically diverse states that combined account for >10% of the entire US population. The SIDs of these states contain unique “visitlink” variables that allow for tracking individual patients over numerous hospital discharges across years.

We used the National Readmissions Database (NRD) (2010-2019) and the 2007-2019 National Inpatient Sample (NIS) to evaluate the generalizability of observed trends to the entire US. The NRD is an all-payer inpatient database designed from 27-30 HCUP partner states and has unique visitlink variables that allow for tracking readmissions within a one calendar-year period. Further details on the NRD are available at https://www.hcup-us.ahrq.gov/nrdoverview.jsp. However, the NRD lacks information on race, therefore, we used the National Inpatient Sample (NIS) to determine if detected trends in racial subgroups extended to the entire country. The NIS is a 20% deidentified stratified random sample of all discharges in the US. Provided weights in the NIS allow for the computation of the entire universe of hospitalizations from the 20% in the NIS. However, index vs follow-up admissions cannot be differentiated.

We conducted this study in accordance with RECORD (**RE**porting of studies **C**onducted using **O**bservational **R**outinely-collected **D**ata) guidelines^10^. FOO was responsible for data stewardship.

### Study Population

We identified all adult discharges (≥18 years) with a primary or secondary diagnosis of IE by querying the SID/NRD/NIS as appropriate using International Classification of Diseases (ICD) Ninth Revision diagnostic codes 421.0, 421.1, 421.9, 112.81, 036.42, 098.84, 424.90, 424.91 and 424.99 before September 2015 and ICD-10 codes I330, I339, I38, I39, A3282, A3951, A5483 and B376 afterwards (Supplemental Table 1). This combination of ICD-9 codes have been validated in multiple studies^2,11^ and found to have high sensitivity (>90%)^2,11^ and positive predictive value (PPV) (80%)^2^ for IE. ICD-10 codes have also been shown to have high accuracy for IE^12^. Study exclusions are depicted in Supplemental Figure 1.

### Definition of incident infective endocarditis

An incident IE case was defined as first discharge for IE without any prior hospitalization for IE in that patient in the preceding years in the SID. We utilized a 2-year look back period (2005-2006) to minimize the influence of old IE discharges on study estimates. Utilization of a 4-year look back period did not significantly affect observed trends.

For the estimation of age and sex-specific national IE incidence counts, we utilized the visitlink variable in the NRD to identify all annual first hospitalizations in the NRD within each calendar year. Because a significant proportion of IE discharges may have carried over or presented for readmission beyond the one calendar-year of possible identification in the NRD, we calculated the proportion of all patients in the SID who had at least one discharge containing codes for IE in a year beyond that of their index admission and deducted a similar proportion from all annual estimates (9.7%) (Supplemental Figure 1).

Furthermore, to obtain race specific estimates, which is not possible with the NRD, we first obtained the weighted annual proportion of first admissions in each of the study’s age/sex categories and then applied the proportion for each year to the weighted number of deidentified admissions contained in the NIS for that year in the corresponding age/sex group. Because the NRD is not available in the years before 2010, we used the age/sex-specific readmission proportion for the 2010 NRD in all NIS years preceding 2010. We also applied the proportion of year-over-year readmissions in the SID to the NIS estimates and deducted a conservative 5% proportion from the estimates to account for possible estimates in non-residents, based on a 3.2% non-residents admission rate in the NRD (Supplemental Figures 2). Utilization of this approach yielded age and sex-specific incidence estimates that were within range of those obtained using the NRD alone. We then stratified the weighted counts into various racial subgroups.

### US Population data

We obtained yearly estimates of the mid-year population of the selected states by age, sex and race for the entire study period from the US Census Bureau website (census.gov/).

### Definition of IE risk factors and covariates

Opioid use disorder was determined using the HCUP constellation of ICD-9 and ICD 10 codes for determining opioid use disorder^13^ and contained in Supplemental table 2. All other covariates were defined using constellation of ICD-9 and ICD-10 codes as contained in Supplemental Table 2. To optimize capture of comorbidities, we identified all prior non-IE related discharges in these patients during the period 2005-2018 and defined these covariates as present if contained in a prior hospital stay.

Race/ethnicity were obtained using the HCUP variable “RACE”and sex data obtained using the variable “FEMALE”. Further details on race/ethnicity in the SID are available at https://hcup-us.ahrq.gov/db/vars/siddistnote.jsp?var=race

### Statistical analysis

Baseline characteristics of patients were presented using descriptive statistics. We evaluated for possible linear trend in the age/sex-specific prevalence of each characteristic over time by using logistic regression models with each risk factor as the dependent variable and year of discharge as the independent variable, evaluated continuously, with significance of differences in trend over time assessed using the Wald test. We utilized additional models with interaction terms for age/sex group and calendar time or race/ethnicity interaction with calendar time to compare the pace of change in prevalence between demographic groups over time.

We combined IE incidence counts with census population at-risk to compute incidence. Before computing incidence, we multiplied obtained counts with the combined PPV of the constellation of ICD-9 codes and ICD-10 codes using estimates from prior validation studies (80% for ICD-9^2^ and 78% for the ICD-10 discharges^12^) to ensure that the final incidence counts reflect the likely proportion of actual cases^14^. We age and sex-standardized overall and race-specific incidence estimates to the 2010 US population and utilized 2-proportions Z-test to test for between group differences in incidence. We fitted join point regression models with permutation model selection method and autocorrelation errors to identify points of change in IE incidence and to quantify the annualized percentage change (APC) in incidence over time.

We further utilized multivariable-adjusted negative binomial regression models adjusted for age, sex and the Exhauster comorbidity score^15^ to compare the prevalence of selected IE risk factors between racial/ethnic subgroups and to determine trends over time.

All primary analyses were done by F.O.O. using Stata 16 (College Station, TX). Incidence computation and join point regression were done by A.A and F.O.O using Microsoft Excel (Microsoft Corp, Redmond, WA) and Join point Software version 4.8.0.1 (National Cancer Institute, Bethesda, Maryland). A 2-tailed α of < 0.05 was required for statistical significance. Because this was a purely descriptive and exploratory study with no specific hypothesis tested, adjustment for multiple comparison is not necessary^16,17^. We considered the weighting, clustering and stratification needed in the NIS and NRD analyses through use of relevant discharge weights and stratification variables. We utilized the NIS “TRENDWT” to account for the NIS redesign in 2012 in all years preceding 2012 as recommended.

### Standard Protocols, Approvals and Data availability

The portion of this manuscript done using the SID was approved by the HCUP after the agency reviewed this project proposal and found it to be consistent with its data use agreement. According to HCUP, utilization of the NIS and NRD does not require approval by an Institutional Review Board. All data used in this study are available for direct purchase from HCUP. The authors are bound by agreement not to directly share HCUP data.

### Missing Data

Race data was missing in 0.6% of patients in the SID and 2.3% of discharges in the NIS. Hospital discharges with missing race data were classified in an unknown category in the SID due to relatively small percentage. Missing race data in the NIS was handled using multiple imputation^18^.

## Results

### Baseline Characteristics and risk factors for IE in the SID and NRD

There were 98,221 incident IE hospitalizations in the combined SIDs of Florida (n=55,735) and New York (n=42,486) across the period 2007-2018. 92.6% of these IE were designated as present-on-admission. 70.0% were NHW patients and 60.6% were ≥ 65 years of age (Supplemental Table 3). However, the proportion in young women 18-44 years more than doubled (4.6% to 10.6%) and that in young men 18-44 years of age increased by >75% (5.6% to 9.4%) over the period 2007-2018, so that the proportion in patients ≥ 65 years declined over time (all *p*-for-trend<0.001) (Supplemental figure 2). 17.6% of all IE patients had drug use disorder (DUD) and specifically, 11.2% had opioid use disorder (OUD). Prevalence of any DUD and OUD was highest in young women 18-44 years and men 18-44 years (p-value for all pairwise comparisons to women 18-44 years <0.001) (Supplemental Table 3). Prevalence of OUD and any DUD increased by >2-fold in all age and sex groups over time but the pace of this increase was fastest in women 18-44 years, followed by men 18-44 years (all p-for-interaction <0.001) (Figure 1). In 2018, 80.4% of women 18-44 years had comorbid DUD.

**Figure 1.**
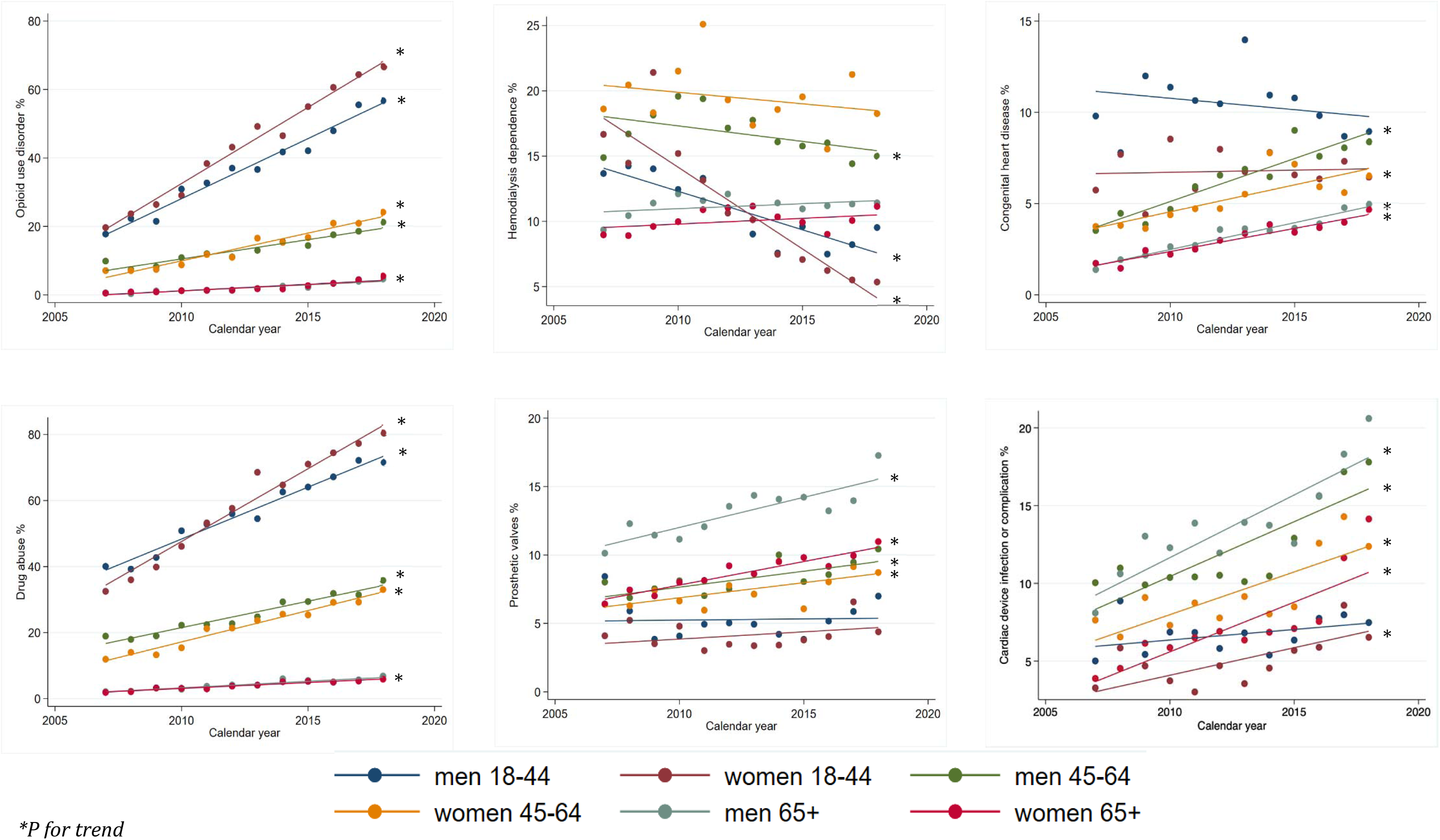
Trend in the prevalence of infective endocarditis predisposing conditions in 98,221 incident infective endocarditis patients in New York and Florida from 2007-2019.

The proportion of IE admissions with implanted cardiac device infections or complication also more than doubled in all age groups over time (all *P* for trend <0.001) except in men 18-44 years (*p=*0.122) but overall prevalence of device infection was highest in elderly men (Figure 1). The proportion of endocarditis in patients with prosthetic valves also increased over time in most age groups in the SID (Figure 1) and in all age groups nationally (Supplemental Figure 3). However, the prevalence of hemodialysis dependence declined in all age groups < 65 nationally (Supplemental Figure 3). Proportion of patients with multiple comorbidities also increased over time.

### Age and sex-specific incidence of IE in cases per 100,000 population

From 2007-2018, the average annual age and sex standardized incidence of IE in the SID was 19.2 (95%CI 18.7-19.6) but there was marked heterogeneity in incidence by age/sex. Age-standardized incidence in men (20.9 [95%CI 20.2-21.7) was 1.2-times that of women (17.5 [95%CI 16.8-18.2]) but on stratification by age, this sex gap in incidence was evident only in age groups 45-64 and ≥65 years (Figure 2).

**Figure 2.**
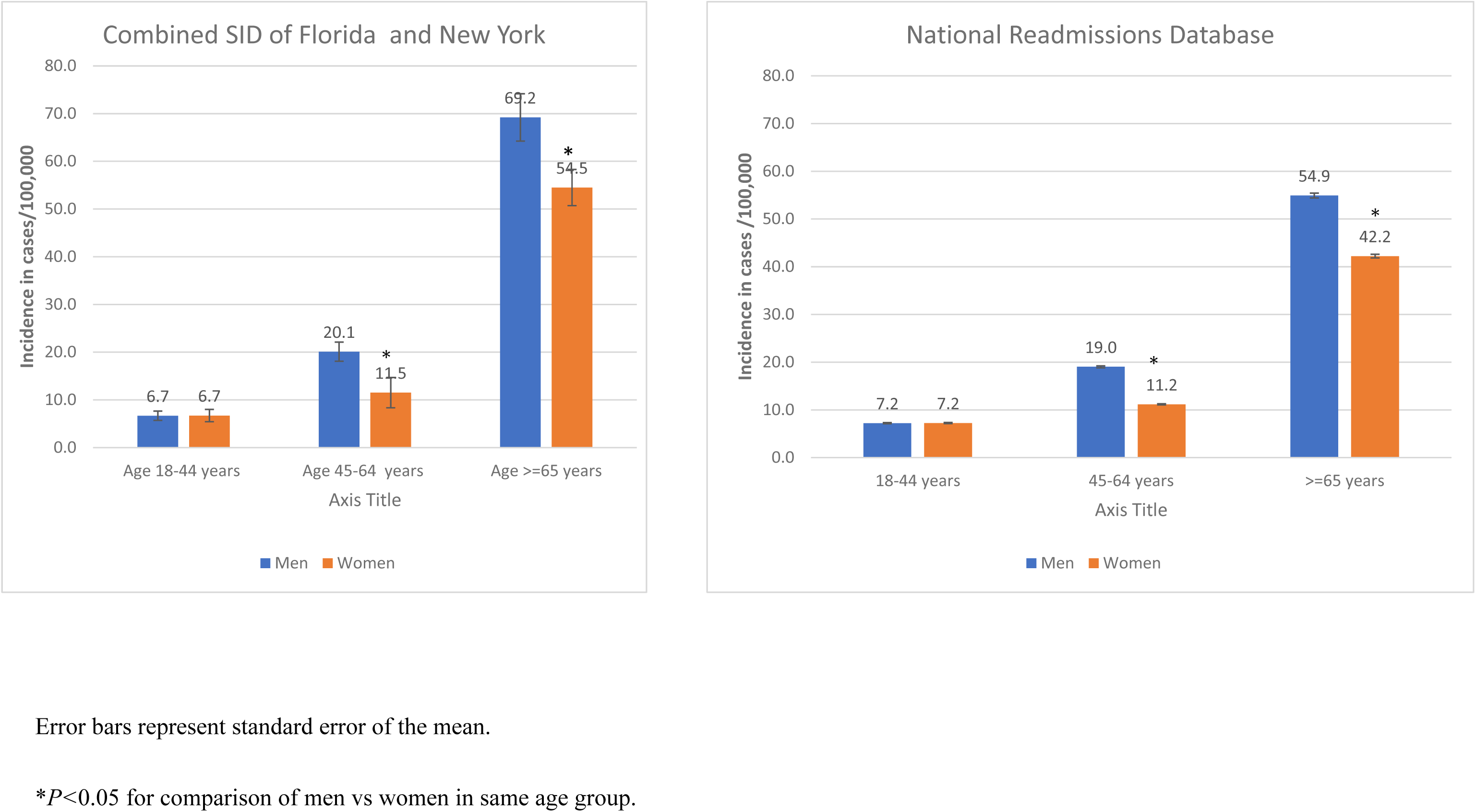
Average annual age-specific incidence of infective endocarditis across the period 2007-2018 in Florida and New York combined and in the entire United States from 2010-2019.

Nationally, the average age and sex-standardized annual burden of first hospitalizations for IE in the NRD was 16.9, (95%CI 16.4-17.5). The age and sex-distribution of these national cases mirrored those of the incident cases in the SID, with age-standardized burden in men (18.6[95%CI 17.8-19.4]) being 1.2 times that of women (15.3 [95%CI 14.6-16.0]) and higher incidence in men compared to women only in age groups 45-64 years and ≥ 65 years (Figure 2).

### Age and sex-specific trends in incidence over time

Across the period 2007-2018, the age- and sex-standardized incidence of IE did not change significantly (APC 0.5%, 95%CI −1.2% to 2.1%) in the SID. However, further stratification by age and sex elucidated important trends. Incidence increased exponentially in women 18-44 years (APC 11.0%, 95%CI 9.3% to 12.7%), men 18-44 years (APC 7.3, 95%CI 6.4 to 8.1%), and 45-64 years (APC 1.5%, 95%CI 0.7% to 2.4%) but declined in women ≥65 years (APC −2.8%, 95%CI −5.5 to −0.0) (Figure 3). Incidence in women 45-64 years and men ≥65 years did not change significantly over time. This increase occurred both in individuals residing in Florida (supplemental figure 4) and those residing in New York (supplemental figure 5)

**Figure 3.**
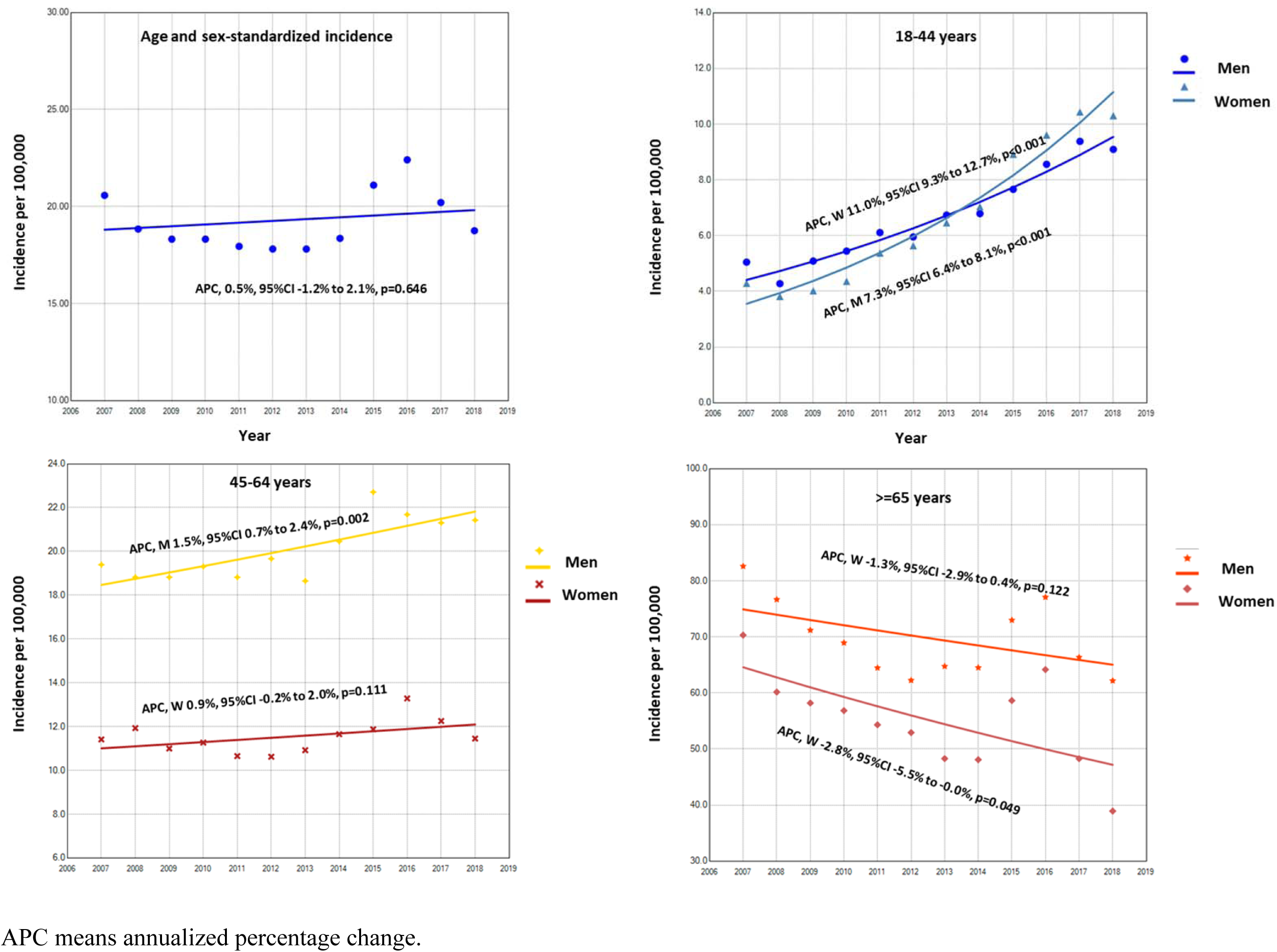
Joinpoint regression of age/sex-standardized and age specific incidence of infective endocarditis in Florida and New York from 2007-2018 according to sex.

Again, the estimated national age and sex-standardized incidence of IE in the NRD also followed similar trends to those in the incidence states, increasing exponentially over time in men 18-44 years, women 18-44 years and men 45-64 years but declining in women ≥65 years (Figure 4).

**Figure 4.**
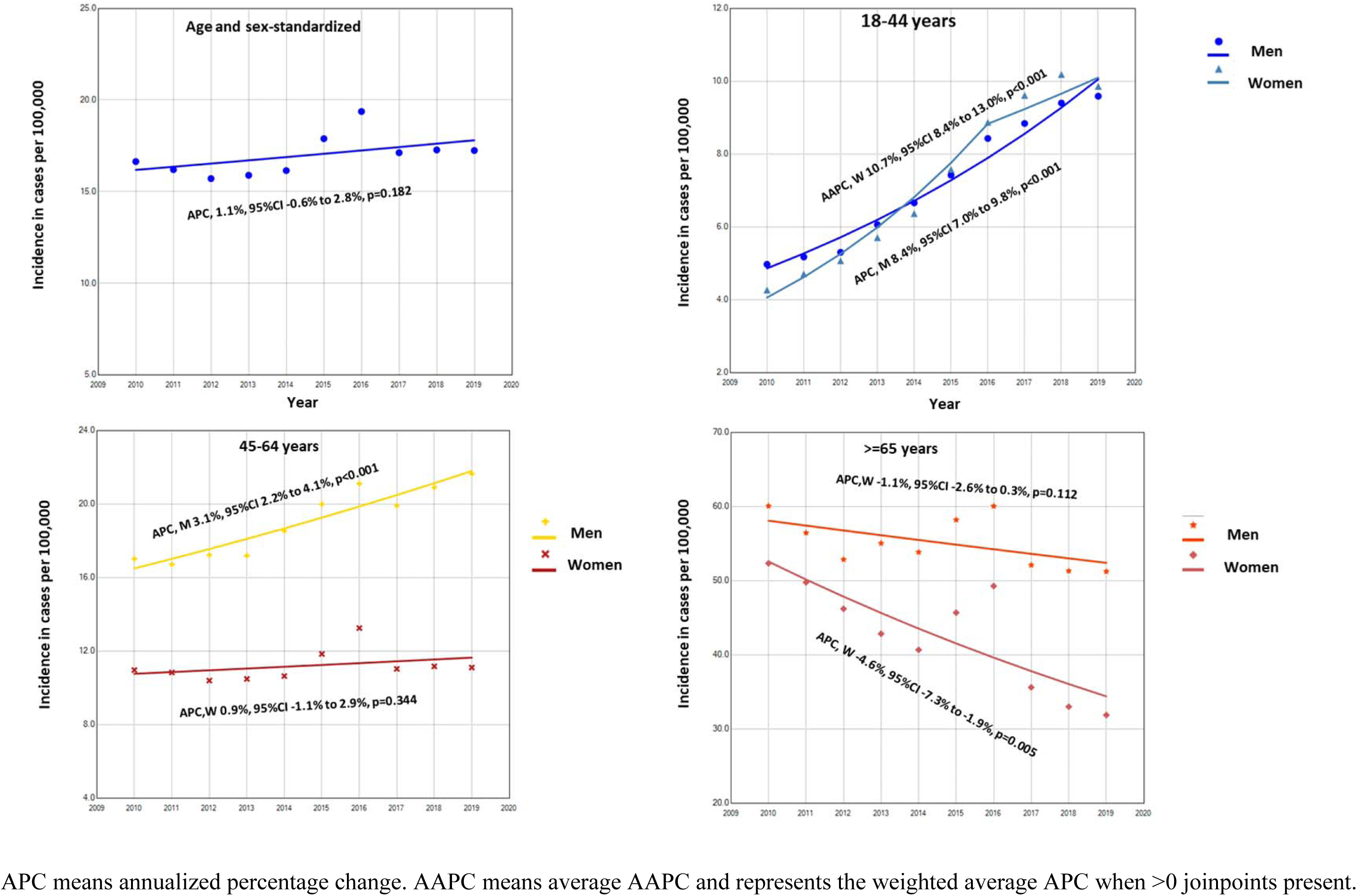
Joinpoint regression of annual age/sex-standardized and age specific first discharges for infective endocarditis in the United States from 2010-2019 according to sex.

### Race-stratified clinical characteristics and multivariable association of demographic characteristics with selected IE predisposing conditions

The mean age of IE was at least 5 years younger in NHB compared to other races (Supplemental Table 4). Approximately 40% (39.5%) of all black patients were 45-64 years old but <30% of all patients in other race/ethnic groups were in this age group (Supplemental Table 4). A greater proportion of NHW had OUD (12.0%) compared to NHB (8.6%) and this proportion increased at a significantly faster pace in NHW compared to other races in all age groups <65 years, particularly among women 18-44 years (Supplemental Figures 6-12). Almost half of all NHB patients (49.8%) had CKD and just over one-third (34.5%) were hemodialysis-dependent and this proportion declined over time in NHB women 18-44 years (supplemental Table 4 and Supplemental Figure 7.

After multivariable adjustment for age, sex, insurance status and comorbidity burden, hemodialysis dependence was >2.5 times more prevalent in NHB vs NHW patients (Prevalence rate ratio (PRR) 2.65, 95%CI:2.54-2.77) but OUD was 43% less prevalent in NHB patients compared to NHW (PRR 0.57, 95%CI 0.54-0.61) (Table 1). In these models, hemodialysis prevalence declined by 2% per unit increase in year (PRR 0.98, 95%CI 0.98-0.99) while those of other conditions including congenital heart disease, history of prosthetic valves, presence of pacemaker and cardiac device infections increased over time. Prevalence of OUD increased by 10% with each unit increase in year after multivariable adjustment (PRR 1.10, 95%CI 1.10-1.11).

**Table 1.** Multivariable comparison of the prevalence of selected predisposing conditions for infective endocarditis between demographic subgroups.

|  | Congenital heart disease |  | Prosthetic heart valves |  | Pacemaker or defibrillator present |  | Hemodialysis dependence |  | Opioid use disorder |  |
| --- | --- | --- | --- | --- | --- | --- | --- | --- | --- | --- |
|  | PRR (95%CI) | P-value | PRR (95%CI) | P-value | PRR (95%CI) | P-value | PRR (95%CI) | P-value | PRR (95%CI) | P-value |
| **Age, years** |  |  |  |  |  |  |  |  |  |  |
| 45-64 vs 18-44 | 0.63 (0.57-0.68) | <0.001 | 1.26 (1.14-1.38) | <0.001 | 1.35 (1.26-1.45) | <0.001 | 0.78 (0.73-0.83) | <0.001 | 0.47 (0.44-0.49) | <0.001 |
| ≥65 vs 18-44 | 0.34 (0.31-0.38) | <0.001 | 1.53 (1.38-1.69) | <0.001 | 1.67 (1.55-1.80) | <0.001 | 0.35 (0.33-0.38) | <0.001 | 0.07 (0.06-0.07) | <0.001 |
| **Women vs men** | 0.83 (0.78-0.88) | <0.001 | 0.71 (0.69-0.75) | <0.001 | 0.67 (0.65-0.69) | <0.001 | 0.92 (0.88-0.95) | <0.001 | 1.08 (1.04-1.12) | <0.001 |
| **Race** |  |  |  |  |  |  |  |  |  |  |
| NHB vs NHW | 0.87 (0.79-0.95) | 0.003 | 0.68 (0.63-0.73) | <0.002 | 1.02 (0.98-1.07) | 0.362 | 2.65 (2.54-2.77) | <0.001 | 0.57 (0.54-0.61) | <0.001 |
| Hispanic vs NHW | 1.05 (0.95-1.16) | 0.333 | 0.93 (0.87-1.00) | 0.043 | 0.99 (0.94-1.04) | 0.714 | 1.76 (1.66-1.86) | <0.001 | 0.77 (0.72-0.82) | <0.001 |
| Asian/Pacific Islander vs NHW | 1.02 (0.80-1.31) | 0.858 | 0.96 (0.80-1.14) | 0.638 | 1.03 (0.91-1.17) | 0.648 | 2.40 (2.11-2.74) | <0.001 | 0.24 (0.17-0.33) | <0.001 |
| Other/unknown vs NHW | 1.04 (0.91-1.18) | 0.579 | 1.03 (0.94-1.13) | 0.511 | 1.08 (1.01-1.16) | 0.020 | 1.80 (1.66-1.94) | <0.001 | 0.77 (0.71-0.84) | <0.001 |
| Year | 1.03 (1.03-1.04) | <0.001 | 1.03 (1.03-1.04) | <0.001 | 1.03 (1.02-1.03) | <0.001 | 0.98 (0.98-0.99) | <0.001 | 1.10 (1.10-1.11) | <0.001 |

Abbreviations. PRR stands for Prevalence rate ratio. NHW means Non-Hispanic White and NHB means Non-Hispanic Black All models also adjusted for insurance status, Elixhauser comorbidity score, hospital state and patient zipcode.

### Race, age and sex-stratified incidence in cases per 100,000 population

The average annual age and sex-standardized incidence of IE was significantly higher in NHB (22.5, 95%CI 21.0-23.9) compared to NHW (20.0, 95%CI 19.3-20.6), Hispanic (13.2, 95%CI 12.3-14.1) and Asian/Pacific Islander individuals (5.9, 95%CI 4.7-7.1) (*p-*value for all comparison to NHB <0.001). This disparity in incidence was driven mainly by an at least ≈2-fold greater IE incidence in NHB in women 45-64 years and no < 50% greater incidence in men 45-64 years compared to those of other racial/ethnic groups (Figure 4). This racial gap in incidence in these middle-aged individuals was consistent across both states (Supplemental Figure 6), however the incidence patterns in other age/sex groups in the two states were very divergent. Whereas incidence was higher in NHB men and women ≥65 years residing in Florida compared to other races, incidence was higher in NHW compared to other races in residents of New York. Among Florida residents and in both states combined, incidence was higher in NHW compared to other races in men and women 18-44 years old (all p-values for pairwise comparisons <0.001) but was higher in NHB compared to other race/ethnic groups in women 18-44 years in New York residents (Figure 5 and supplemental figure 6).

**Figure 5.**
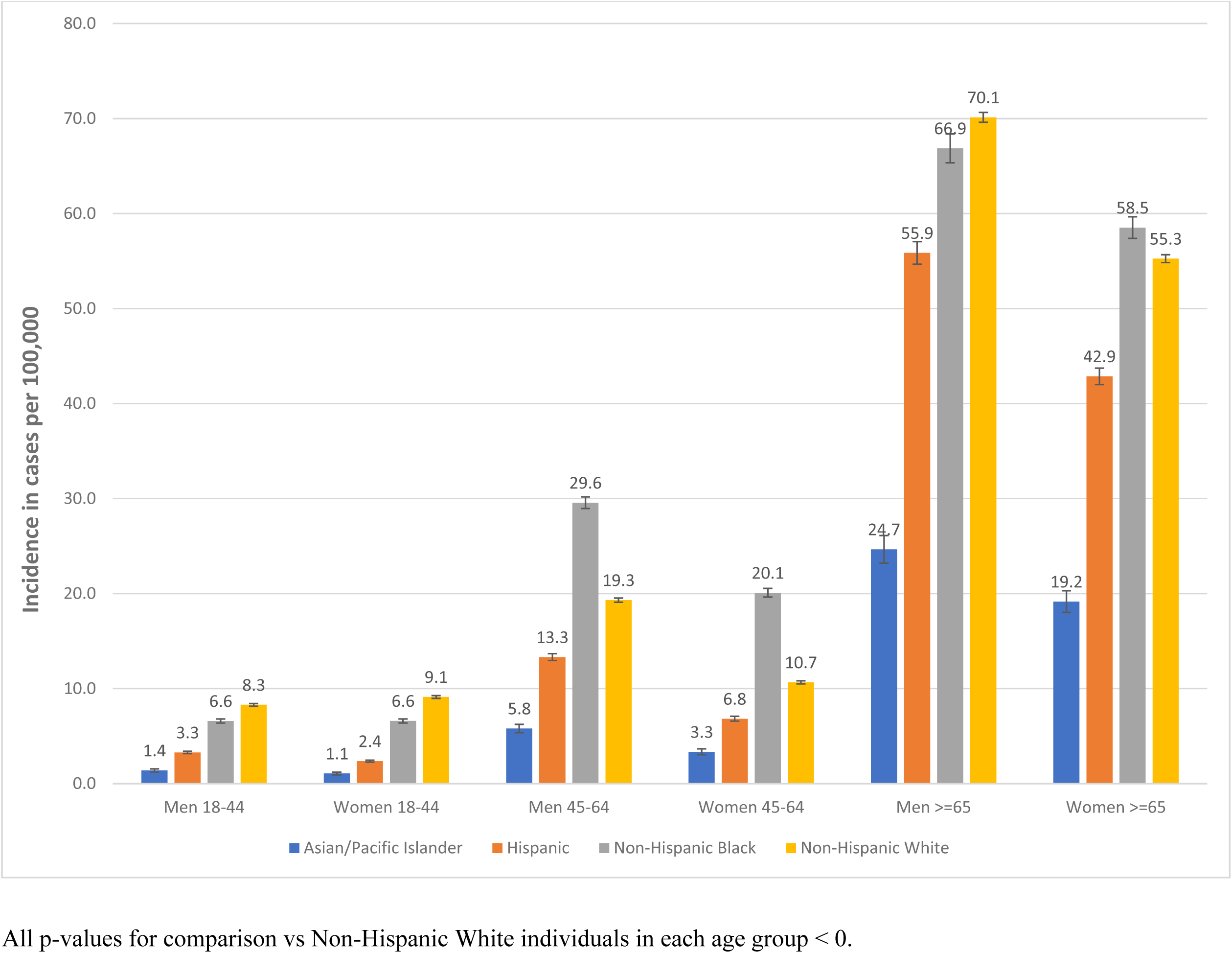
Racial differences in the average annual incidence of infective endocarditis from 2007-2018 in Florida and New York combined in various age and sex groups.

### Trends in age and sex-specific incidence stratified by race

IE incidence increased exponentially in NHW men (APC 10.6%, 95%CI 9.8% to 11.8%) and women (APC 16.6%, 95%CI 14.9% to 18.4%) 18-44 years such that by 2018, incidence has increased by >2-fold in men and by >4-fold in women in this racial subgroup compared to 2007 (Figure 6). Incidence in Hispanic men 18-44 yo also increased by 5.6% annually (APC 5.6%, 95%CI 4.1% to 7.1%) but pairwise comparison of the regression mean functions for NHW vs Hispanic indicated that the pace of increase was slower in Hispanic vs NHW men ((*p* < 0.001). Interestingly, incidence did not change significantly in NHB men or women 18-44 or 45-64 years old in either state (Figure 6). Similarly, incidence increased marginally in NHW men (APC 2.8%, 95%CI 1.6% to 3.9%) and NHW women (APC 1.8% (95%CI 0.2% to 3.4%) 45-64 but did not change significantly in men and women in other racial/ethnic groups in this age group (figure 6). These findings occurred both in New York and in Florida (Supplemental figures 14 and 15). In contrast, within the ≥65 year age group, the incidence declined significantly in NHB women and Asian Pacific Islander women and showed a trend towards significant decline in NHW women (p=0.063) Figure 6.

**Figure 6.**
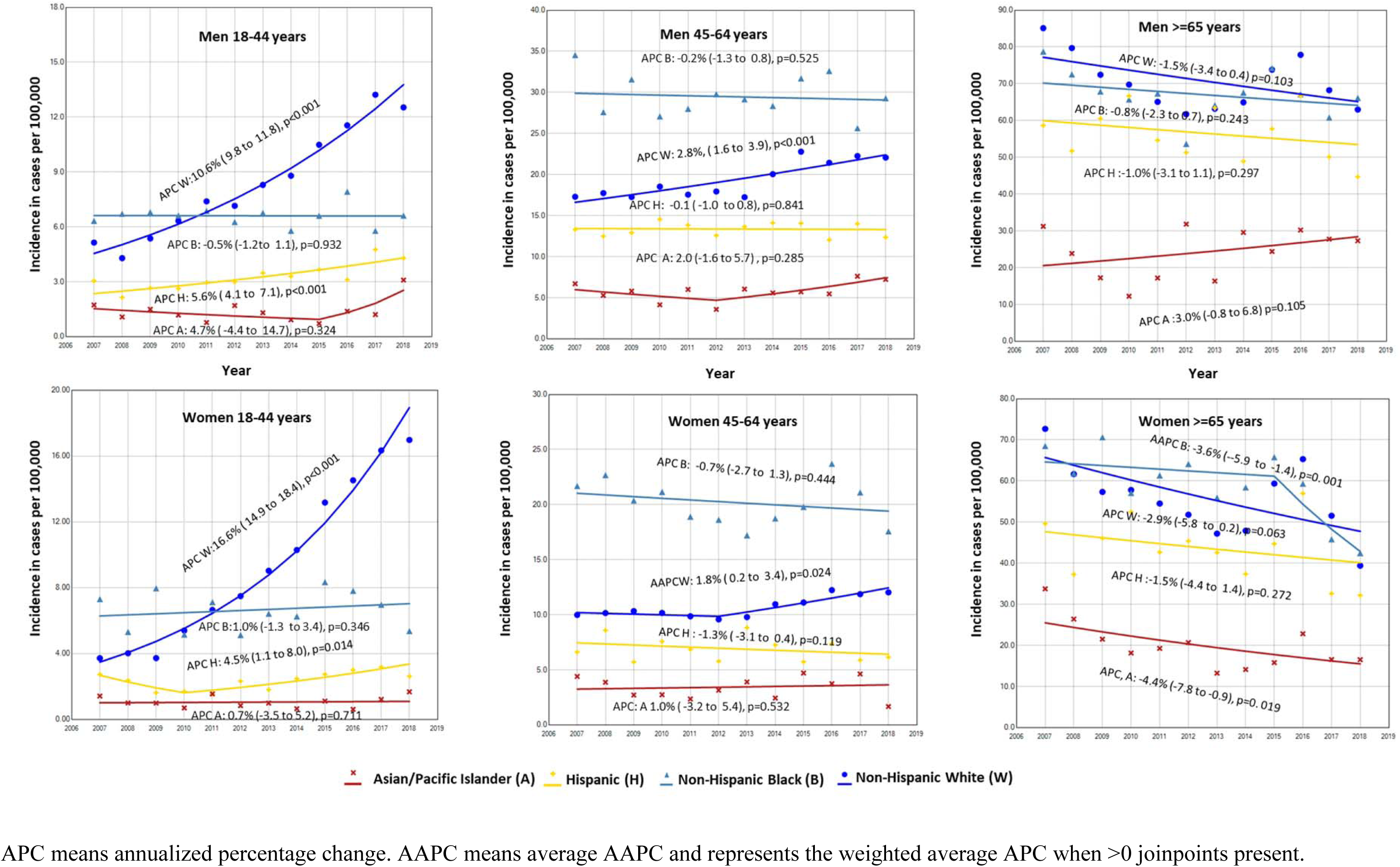
Joinpoint regression of age specific incidence of infective endocarditis in Florida and New York according to race in various age and sex groups.

After applying NRD readmission proportions and SID year-to-year readmission proportions to the NIS, the estimated burden of IE nationally in NHW women 18-44 years increased exponentially by 17.9% annually (APC:17.9% [95%CI 15.7 to 20.1]) such that by 2019 incidence has increased by >4-times that of 2010 (Figure 7). That of NHB women in the same age group remained unchanged over time (APC: 1.0% [−0.8 to 2.9]). Burden increased by 13.2% annually in NHW men 18-44 years (APC W:13.2% [95%CI 12.4 to14.1]) and by 6.5% in Hispanic patients (APC: 6,5% [95%CI 4.9 to 8.1]). However, in contrast to the SID, burden also increased in NHB men 18-44 (APC:4.4% [95%CI 2.7 to 6.2]) and 45-64 years old (APC: 1.5% [95%CI 0.7 to 2.3]) (Figure 7).

**Figure 7.**
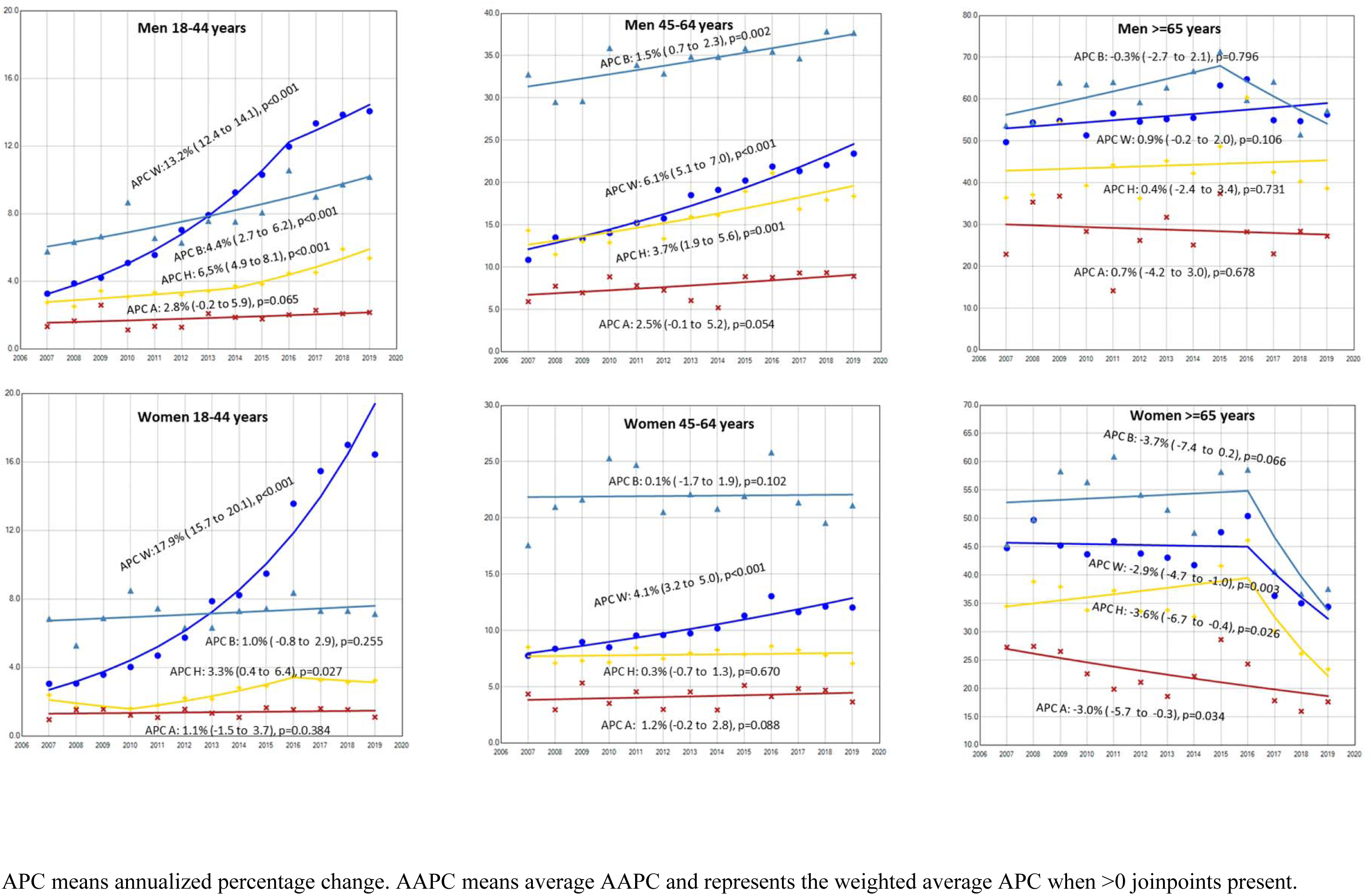
Joinpoint regression of race specific discharges for infective endocarditis in the United States from 2007-2018 according to age and sex group categories.

## Discussion

In this contemporary analysis of the SID, we highlight critical trends in the incidence of IE in the states of New York and Florida over the period 2007-2018. We found that the combined age and sex-adjusted incidence of IE in both states was approximately 19 cases per 100,000 population but incidence differed markedly by age, sex and race. Incidence increased with age in both sexes and across all races, but the age-standardized incidence was higher in NHB compared to other races. The age and sex-standardized incidence of IE did not change significantly over the study period but incidence in women and men 18-44 years old increased exponentially, while that of men 45-64 years old increased marginally over time. A disproportionately greater portion of this increase occurred in NHW women and men 18-44 years and in NHW men 45-64 years old.

Prior population-based studies evaluating the incidence of IE in the US over the last two decades have reported varying estimates ranging from 7 to 290 cases per 100,000 population^2,11, 19–25^. However a significant proportion of these studies were serial cross-sectional studies done solely using national^19,22,25^ or other state^4^ administrative claims databases in which differentiation of index vs follow-up admissions was not possible. Such databases may not be optimal for determining true IE incidence due to very high readmission rate (26.5% of all admissions in the SID portion of our study). Other studies that utilized longitudinally collected data such as Medicare fee-for-service data were restricted to elderly patients^20,21^ or excluded key subgroups of IE patients such as those with implanted electronic-device related infections^23,24^. Remarkably, one study also done using the SID of New York and California, reported age-standardized incidence of 7.6-7.8 cases/100,000 population across the period 1998-2013^2^ which is markedly lower compared to ours. The disparate estimates are partly due to differences in the population studied. While our study focused on adult population only, this study incorporated patients in the entire age spectrum, from neonates to old age, in their age-standardized estimates^2^. Therefore, the considerably lower incidence of infective endocarditis in children in their study^2^ may have skewed their estimates lower. More importantly, methodological differences between both studies were also contributory. The latter study excluded all patients with ICD-9 codes for “Endocarditis valve unspecified” corresponding to ICD-9 codes in the range of 424.9x, because inclusion of these codes resulted in a drop in the PPV of IE (compared to chart review) from 93.9% to 80.1%^2^. However, these codes were present in 45.1% of all endocarditis cases from 2007-2014 in our patient population. We chose to include discharges with these codes in order not to underestimate the true IE incidence burden and also to allow for consistent IE definition between ICD-9 and ICD-10 codes across the study period. Knowing that this approach will increase sensitivity for IE detection but will potentially diminish specificity, we optimized the accuracy of these codes by adjusting incident counts to the combined probability that a patient with these codes had true IE (i.e., the PPV of the codes)^14^. Our age-specific estimates in elderly patients ≥65 years are within range of those of other studies that incorporated these codes in elderly patients^20^. Nonetheless, our findings also agree with those of the SID study^2^ and those of others^11^ reporting no significant increase in age and sex-standardized incidence of IE following the American Heart Association 2007 guideline on antibiotics prophylaxis for endocarditis^1^. We however extend the results of these studies by uncovering unique patterns in age-specific incidence over time that firstly are indicative of a demographic shift in IE incidence in the US towards younger adults and secondly highlight key racial differences in age-specific trends that have tremendous public health and policy implications.

The dramatic increase in incidence in young men and women is likely due in large part to the prevailing trends in the OUD epidemic in the population. Prevalence of implanted device-associated infections/complications also increased over time in these young women in the SID and in both sexes nationally, but the increased incidence risks attributable to this condition is likely smaller compared to OUD due to qualitatively higher overall OUD prevalence. In this study, the prevalence of DUD and OUD more than doubled over time in all demographic subgroups over time, but the young women/men 18-44 years who experienced the greatest increase in prevalence of OUD and DUD over time, also had the greatest increase in incident IE. The disproportionate increase in OUD prevalence in young NHW women and men also corresponded very well with the excess increased incidence noted in this racial subgroup compared to other races. In the National Survey on Drug Use and Health from 1999-2018, the adjusted relative risk of past-year prescription opioid misuse was 19%-41% less in NHB, 13%-35% less in Hispanic and 52-67% less in Asian people compared to NHW^26^. This racial disparity in OUD was even more prominent for heroin use, where other races had 47%-65% lower adjusted relative risk of past-year heroin use compared to NHW at every period from 1999-2018. The age-adjusted prevalence of past-year heroin use more than doubled in NHW and also increased at a faster pace in NHW people compared to all other races across the period 1999-2018, a period that encompasses that of our current study^26^.

It is noteworthy that the OUD prevalence also increased in other racial/ethnic subgroups of this study albeit at a slower pace compared to NHW so this increased burden of OUD in IE is not isolated to NHW people. Whereas in the SID of New York and Florida, the increased OUD prevalence translated to higher overall IE incidence in Hispanic patients 18-44 years old, the incidence in NHB remained unchanged in these age groups. The prevalence trends for some of other IE risk factors in our study population (including declining hemodialysis-dependence and HIV/AIDS prevalence) which are differentially more prevalent in young NHB, suggests that the increase in OUD-associated cases in the NHB subgroup may have been counterbalanced to some degree by declining IE incidence in patients with some of these conditions. However no definite inferences can be deduced from our retrospective analysis of a claims database. In fact, in our nationally projected estimates, the incidence of IE also increased in NHB men 18-44 and 45-64 years of age as well.

Regardless of the etiology, exponentially rising OUD-associated IE incidence in young patients implies that current measures to reduce IE risk in PWID are less than satisfactory. These findings are also a call to policy makers to intensify education for safer injection practices to PWID in these young age groups^28^. Perhaps legislation to amplify proven harm reduction strategies such as syringe services programs^29^, which currently may not be legally available in all states^28^ may also be needed.

That the incidence of IE should be higher in NHB compared to NHW is expected. Poorly controlled hypertension, chronic kidney disease and eventual hemodialysis dependence are more prevalent in NHB compared to other races^30^. Conditions such as HIV/AIDS which may increase susceptibility to IE are more prevalent in NHB people in the US and in our study population^30^. Inflammatory diseases such as systemic lupus erythematosus or sarcoidosis which may necessitate immunosuppressive therapy are also more prevalent and notably more severe in NHB in the general population compared to NHW^31,32^. These racial disparities are possibly rooted in or perpetuated by socioeconomic inequity and structural racism^32,33^. However, to the best of our knowledge, this is the first study to evaluate incidence of IE according to age and sex in various racial/ethnic subgroups in the US. Whereas Toyoda et al, reported no change in IE incidence in NHW and non-white patients across the period 1998-2013^2^, the magnitude and direction of differences in race-specific incidence estimates were not formally quantified. Incidence of IE has been shown to be higher in NHB Medicare beneficiaries ≥ 65 years compared to NHW over the period 1999-2010^20^ but studies specifically quantifying IE incidence according to race in other age groups, especially according to sex groups of the population are notably lacking. In this study we demonstrate that the age- and sex-standardized incidence of IE in NHB is 1.2-times that of NHW, 1.7-times that of Hispanic and >3-times that of Asian patients. We also demonstrate that as a result of the changing demographic profile of IE over the study period, the higher incidence in NHB may not be present across the entire age spectrum. The only age group where incidence may still be consistently higher in NHB compared to NHW men and women are in those 45-64 years of age. The combined average annual incidence was greater in NHW men and women compared to NHB in patients 18-44 yo. Incidence is higher in NHW vs NHB men ≥ 65 years but lower in NHW vs NHB women ≥65 years. These racial, age and sex differences were however very heterogenous between states. Geographical variabilities in incident IE have been previously described^25^ but it will be of epidemiological interest to see how these racial differences continue to evolve with risk factor changes over time.

Globally, our estimated national incident estimate of IE in the NRD of 18.6 for men and 15.3 for women puts the incidence of IE in the US in the range of those of other high income countries including Australia, Belgium, Denmark, Germany, the Netherlands, Norway and Spain^34^. Our results show that previously described decline in the incidence of IE in older adults ≥65 in the US over the period 2004-2010^20^ have continued into most of the last decade. Our finding of increasing device-associated infections agrees with those from Scotland showing increasing proportion of device-associated infection in Scotland^7^. However, in contrast to this Scottish study that also showed doubling of IE incidence in very elderly patients over the period 1990-2014^7^, overall incidence in elderly women in the US may actually be declining.

### Strength and Limitations

Key strengths of this study include our large sample size that allowed for stratified analysis by key demographic variables which helped us to delineate the specific racial, age and sex groups where IE incidence may be of growing concern. Through our state-specific estimates we highlighted demographic subgroups in which disparities may be consistent across states and others where incidence may differ between states. We also applied novel approaches to obtain national estimates from the NRD and NIS that illustrated that the findings from these two states are likely generalizable to the entire US.

This study should also be viewed in the context of its limitations. Even though we have adjusted our incidence estimates to the estimated PPV of the ICD-9 and ICD-10 codes used in this study, we cannot exclude coding inaccuracies. Our study of trends over time is dependent on the implicit assumption that coding and clinical practices remained unchanged over time. However coding practices is likely to have improved over time. With advances in clinical knowledge over time, index of suspicion for potential IE cases may have increased over time, and with advances in cardiovascular imaging, clinical detection of IE is also likely to have improved with time. This study likely underestimates the true burden of IE in the population as some patients may die before presentation and others may remain undiagnosed even following hospital presentation. Our national estimates, particularly that done using the NIS should be interpreted with caution as it fundamentally assumes that readmission rate is consistent across all races. However, readmission patterns for IE may differ between racial subgroups and this may have marginally biased racial estimates. However, it is likely that racial differences in readmissions may exist. The potential impact of these differences on study estimates is likely minimal. Information on race/ethnicity in the SID is not standardized across states and not always based on the gold-standard of self-report. We have no information on non-binary gender expressions as these are reported as missing in the SID.

## Conclusion

IE incidence more than doubled in NHW men and increased by >4-fold in NHW women aged 18-44 across the period 2007-2018 in New York and Florida. Incidence also increased in Hispanic men 18-44, and in NHW and Hispanic men 45-64 years, but that of NHB did not change significantly over time in these age groups. OUD prevalence trends in IE patients in these racial subgroups suggest that the continued rise in OUD prevalence in the general population may be the primary driver underlying this increase. Intensified educational and policy efforts targeted towards harm reduction in OUD patients are needed to stem this rise particularly in young NHW women and men.

## Figure Legend

Error bars represent standard error of the mean.

## Author Contributions

Dr Otite had full access to all the study data and takes responsibility for the integrity and accuracy of the data analysis.

Study concept and design: Anuforo, Otite

Acquisition, analysis, or interpretation of data: Anuforo, Otite Drafting of the manuscript: Otite

Critical revision of the manuscript for important intellectual content: All authors.

Statistical analysis: Anuforo, Otite.

Administrative, technical, or material support: Anikpezie, Otite

Study supervision: Otite

## Conflict of Interest Disclosures

Dr Chaturvedi is an associate editor for the *Stroke* journal and Dr Ovbiagele is Editor-in-Chief of the *Journal of the American Heart Association.* The other authors have no relevant disclosures.

## Funding source

None

## Data Availability

All datasets used in this study can be obtained from the Healthcare Cost and utilization project

